# Fear and death anxiety among Latin American doctors during the Covid-19 pandemic

**DOI:** 10.1101/2021.02.12.21251445

**Authors:** Sonia Indacochea-Cáceda, Jenny Raquel Torres-Malca, Víctor Juan Vera-Ponce, Miguel A. Pérez, Jhony A. De La Cruz-Vargas

## Abstract

**Introduction:** A patient’s death elicits various feelings arise in doctors such as impotence and guilt that could trigger physical or psychological symptoms. Despite its impact, few studies have explored anxiety and fear of death among physicians.

**Objective:** To evaluate perceptions and responses to one’s mortality among Latin American physicians treating COVID-19 patients.

**Methodology:** This cross-sectional study utilized the Collet-Lester Modified Fear of Death Scale and the Death Anxiety Scale (DAS) to collect data from physicians in Latin America.

**Results:** 219 doctors from Peru, Mexico, Argentina, Colombia, and Bolivia filled out the online questionnaire. Fear of death among the sample population ranged from 56.2% to 90%. Furthermore, the prevalence of “High Anxiety” was 80.8%. A statistically significant association was found between fear according to age and time of graduation (p = 0.010 and p = 0.020, respectively). No differences were found by gender, age, and country of origin or practice.

**Conclusion:** Physicians experience feelings of helplessness and guilt upon the death of a patient and those feelings can trigger physical or psychological symptoms. For these states of fear and anxiety about death, more frequent in times of pandemic, doctors must be prepared, and institutions must provide them with the necessary means to help them overcome these difficult times.

## Introduction

Acceptance of the universality of death, especially one’s own mortality, is a difficult concept for individuals in Western societies who often use euphemisms to describe this inevitable part of human existence. From a clinical perspective, death is defined as “… the irreversible cessation of circulatory and respiratory functions, or the irreversible cessation of all functions of the entire brain…”(1,2). More recently, the Harvard definition(3) of coma or brain death included “unresponsiveness, lack of response, absence of movement or breathing, absence of reflexes with greater delineation of brainstem reflexes and a flat EEG (repeated after 24 hours with no change).

A variety of feelings and emotions are related to the end of life (death and dying) and these reactions are usually dictated by the very culture that prescribes the reactions and undoubtedly affects the person who faces them(4,5). Among the most common emotional responses to death and agony are anxiety, fear, sadness, pain, and depression(6). Ramos and colleagues point out that anxiety and fear are often designed to provide the body with energy to invalidate or counteract the threat through an appropriate response(7) in this case the loss of a person close to the individual experiencing the emotion.

As indicated above, attitudes regarding end of life are generated individually based on the interpretation and interaction with the environment in which they develop. They are, therefore, linked to a personal vision of the world and are also related previous experiences, life expectancy, religious beliefs and perceived control.

Meza(7) and Guinot(8) have concluded that individuals have yet to overcome the universal fear of death, which is related to the existential anguish regarding the end of our days, a situation that worsens with the passing of the years, or when long term care is required. Several models have been developed to understand the process used by humans to cope with death and agony (8–10), among the most often cited being that of Kubler-Ross, who concluded that individuals experience not only fear of death, but also anxiety associated with end of life. According to Kubler-Ross(11) “death remains a terrible and terrifying event, and the fear of death is a universal fear, although we believe that we have mastered it on many levels”(1). Subsequent research in this area has resulted in the concept of death anxiety(12,13).

Limonero described death anxiety as “an emotional reaction produced by the perception of signs of danger or threat to one’s existence”(14). Some authors have described death anxiety as a state of horror: “The horror of death is, then, the emotion, the feeling or the awareness of the loss of one’s own individuality. Emotion, shock of pain, of terror, of horror. Feeling for a break, a bad, a disaster, that is, a traumatic feeling. Consciousness, in short, of a void, of a nothingness, which appears where individual fullness was before, that is, traumatic consciousness”(15). It is the inherent lack of information and control over death that contributes to the fear experienced by individuals.

Mantegazza concluded that death anxiety is present not only in patients close to dying, but also among the staff who attends them(16). For instance, several studies point to the impact of death on nurses who presented anxiety during the dying process of their patients and who experienced fear related to their own mortality(17–20). This fear is the result of the patient’s loss of life and also because of the threat it represents to the health care provider. The impact of this fear of death is amplified when the health care provider does not receive the training and support to deal with death(18,21).

The literature suggests, that patients experience the greatest anxiety following diagnosis, when the prognosis of his disease is bad, or we are in constant uncertainty. Medical professionals on the other hand experience anxiety when they believe they are not capable of facing the suffering and death experienced by their patient or when they do not know some aspect of their disease.

Medical personnel, including physicians, are trained to relieve pain and prolong life. As a result, doctors who face death on a regular basis may have developed particular views and feelings related to this topic. For many clinicians, discussing death and end-of-life with patients and family members can be both a burden and a challenge(12,22–24). In some cases fear of death, feelings of inadequacy, and frustration may lead the doctor to avoid close contact with the patient(25–27,21,20).

Yet, despite the fact physicians are likely to face death as part of their professional work, little or no provision is made in medical schools to care for and understand the non-clinical aspect of death. In fact, many physicians have not received training on how to deal with the issue of death in their patients or on their own, which has led some to call for the inclusion of this type of training in the curricula of medical schools(28).

The global COVID-19 pandemic has forced doctors to deal with the death of both their patients and their peers(9). However, little research has been done to explore these issues among clinicians, especially in Latin America. This study assesses fear and death anxiety among Latin American doctors.

## Methodology

### Study design

This is a cross-sectional study carried out through an online survey distributed four to five months into the global COVID-19 pandemic.

### Subjects

The non-probabilistic sample consisted of medical doctors representing a variety of specialties in the field. Inclusion criteria involved graduation from a medical school and willingness to complete the survey. Those who did not complete the variables of interest for the study were excluded.

### Procedures

The research group was made up of doctors, psychologists and university professors. Demographic variables included gender (male and female); age; time from graduation; country of origin; worksite type (public and/or private institution, retired); specialty (e.g., internal medicine, pulmonology, emergency medicine). A question also measured the number COVID-19 patients treated by the physician.

Data about feelings related to death were collected using the Collet-Lester Modified Fear of Death Scale which consists of 20 items assessing four subscales that include concern or fear for one’s own death, concerns about the death of others, and concerns about the process of dying. This scale has been found to be reliable in each of its subscales.

The Death Anxiety Scale (DAS) which consists of 15 dichotomous statements (True - False) was also used in this study. It is specified that higher scores on the scale show a higher level of anxiety before death. The reliability of the scale is 0.83.

Data were collected in an online format (Google Forms) since it was not feasible to physically recruit the participants due to geographic distribution and social isolation measures. The questionnaire was distributed during the time frame identified above.

### Statistical analysis

Measures of central tendency were estimated for the categorical variables, and in the case of numerical variables, the normal distribution was evaluated and then the best measure of central tendency and dispersion was reported. For the bivariate analysis, the chi-square test or the Student’s t-test was used. Statistical significance was determined at the .05 level with confidence internals of 95%.

Two Poisson GLMs (generalized linear models) based on logistical regression were constructed (crude rates and adjusted). In both models, the measure of association used was the prevalence ratio (PR) using a 95% confidence interval.

### Ethical Considerations

The study was approved by the Human Subjects Committee at the Universidad Ricardo Palma. Passive informed consent was obtained from all the participants who agreed to participate in the study by filling in the virtual questionnaire.

## Results

Two hundred and nineteen, out of 250 physicians, completed the survey. The mean age in the sample was 49.90 (± 11.91), with internal medicine being the highest represented specialty (38.73%). Almost half of the respondents were employed in public institutions (43.84%) and over half resided in Peru (51.60%). The majority of sample respondents (68.95%) indicated treating COVID-19 patients. Table 1 shows the distribution of the sample according to the treatment of patients with COVID-19.

**Table 1.** Descriptive analysis of Latin American doctors during the Covid-19 pandemic.

| Category | n (%) |
| --- | --- |
| **Sex** |  |
| Male | 114 (52.05) |
| Female | 105 (47.95) |
| **Age\*** | 49.90 ($\pm 11.91$) |
| **Especialty** |  |
| Pneumology | 3 (1.37) |
| Internal Medicine | 87 (38.73) |
| Intensive Care | 7 (3.20) |
| Infectology | 5 (2.28) |
| Emergency and Disaster Medicine | 11 (5.02) |
| Other Clinical Specialties | 55 (25.11) |
| Other surgical specialties | 22 (10.05) |
| OBGYN | 9 (4.11) |
| Pediatrics | 20 (9.13) |
| **Employment Site** |  |
| Front Line | 113 (51.60) |
| Other | 106 (48.40) |
| **Time since Graduation** | 23.01 (±11.66) años |
| **Country** |  |
| Perú | 113 (51.60) |
| Other countries** | 106 (48.40) |
| **Treated COVID Patients** |  |
| No | 68 (31.05) |
| Yes | 151 (68.95) |
| **Employment** |  |
| Public Institution | 96 (43.84) |
| Private Institution | 46 (21) |
| Both private and private institution | 73 (33.33) |
| Retired | 4 (1.83) |
| **Scales** |  |
| **Fear Own Death** |  |
| Low | 96 (43.8) |
| High | 123 (56.2) |
| **Fear Process own Death** |  |
| Low | 45 (20.5) |
| High | 174 (79.5) |
| **Fear for Other's Death** |  |
| Low | 21 (9.6) |
| High | 198 (90.4) |
| **Fear Process Other's Death** |  |
| Low | 22 (10) |
| High | 197 (90) |
| **Death Anxiety** |  |
| Low | 42 (19.2) |
| High | 177 (80.8) |

\* Median and SD. \*\* Other Countries: México, Argentina, Colombia y Bolivia.

Findings from this study found death anxiety in three fourths of the sample respondents (81%). Other findings suggest that fear for one’s own death was present in over half of the study respondents (56%) and 80% were concerned about the process of their own death. Sample respondents also express a high level of concern related to the death of others (90%), and about the process of death of others (81%).

Table 2 shows a bivariate analysis of each item of the fear scale including the anxiety scale. A statistically significant association was found in fear of other’s death according to age and time from graduation (p = 0.010 and p = 0.020, respectively). There were no other statistical significant variables.

**Table 2.**
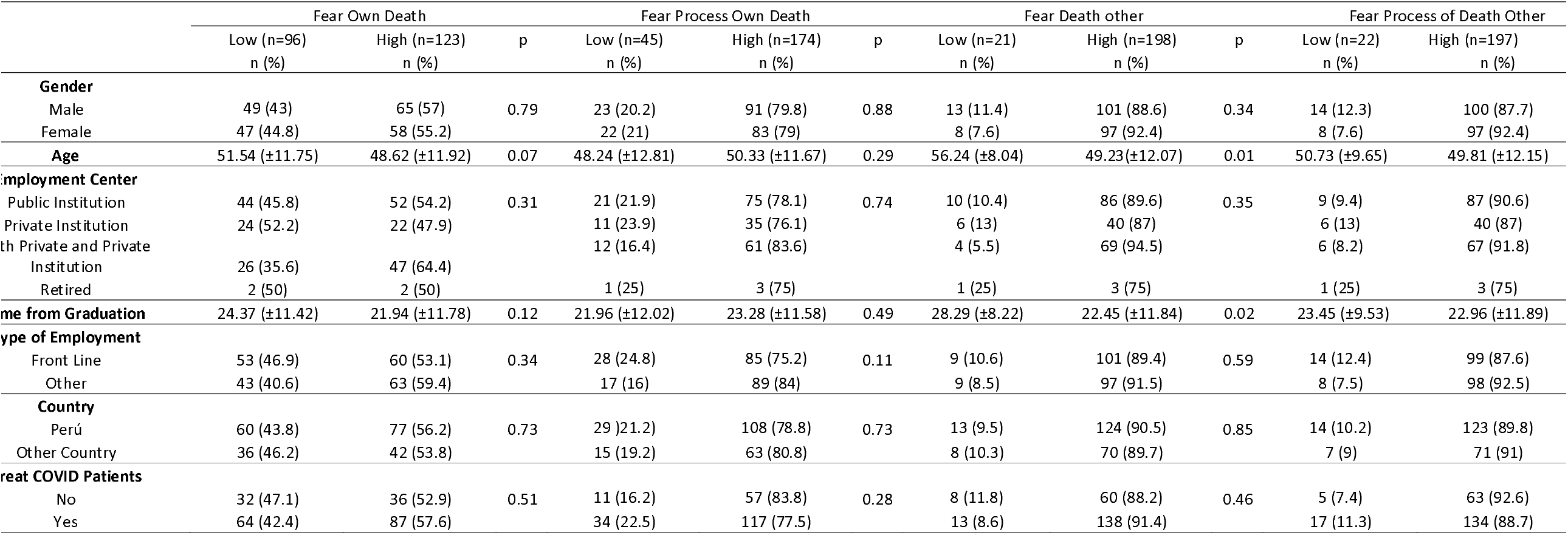
Bivariate analysis of Latin American doctors during the Covid-19 pandemic regarding anxiety and fear of death.

Table 3 shows results from the multiple regression analysis. While the bivariate analysis found a correlation between fear of one’s death and the variables of age and time from graduation, no such correlation was found in the multiple regression analysis. Similarly, no association was observed between any of the variables in the multiple regression analysis.

**Table 3.**
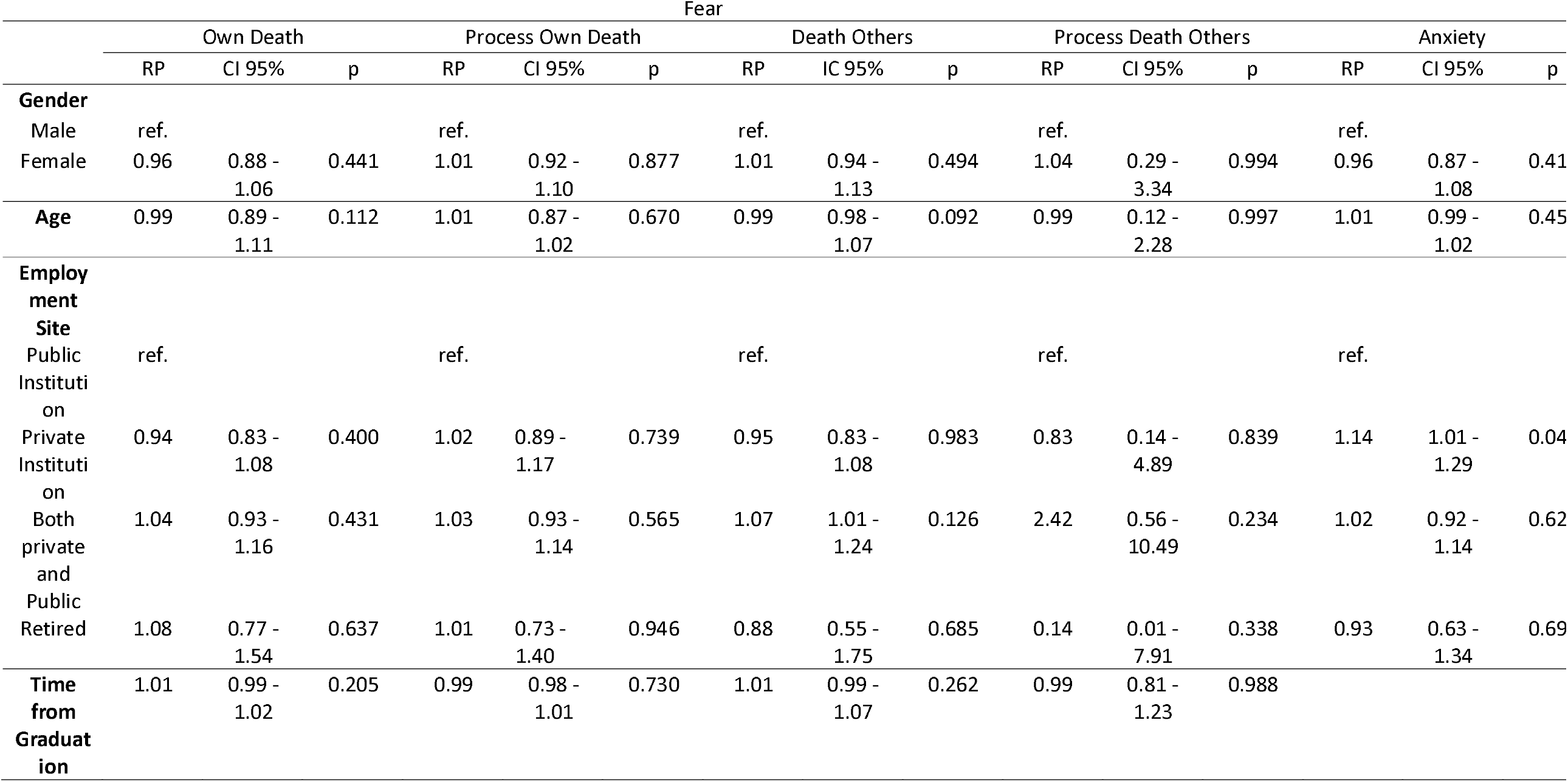
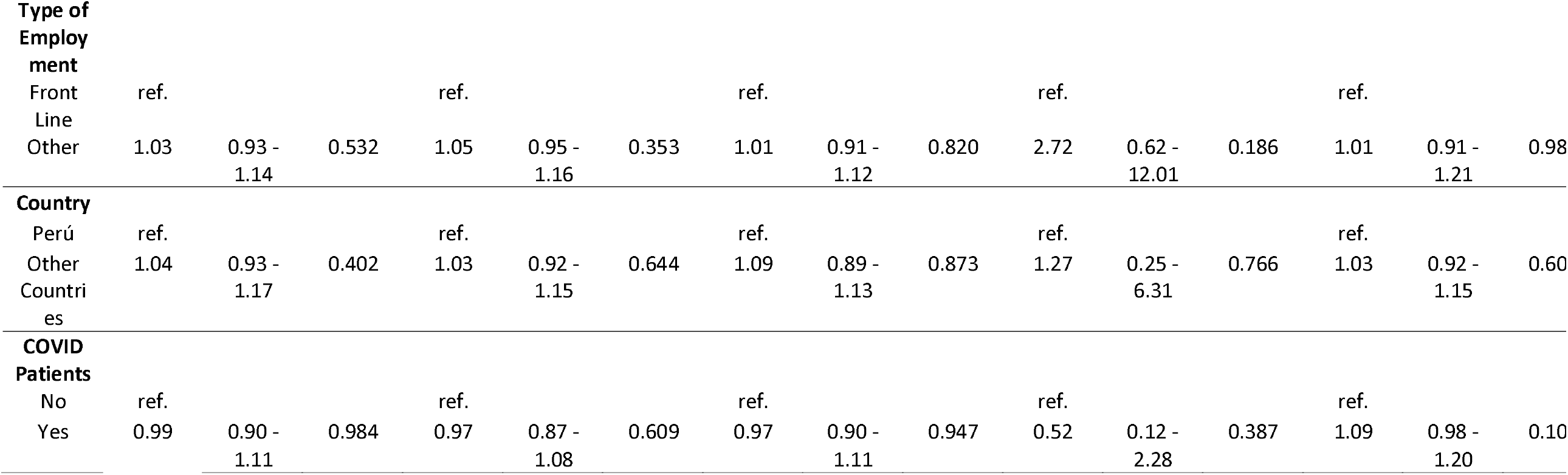
Multiple regression analysis of Latin American doctors during the Covid-19 pandemic regarding anxiety and fear of death.

## Discussion

Death is a traumatic experience that brings fear, anguish, suffering and pain to those who face it and to their survivors(2). The psychological trauma may be felt more acutely by health professionals due to the death of patients under their care. Feelings of anxiety, depression, insomnia and stress may be exacerbated through the risk they face to their own health and the fear of propagation of disease in their immediate environment(26). Emerging literature suggests that this is true of health professionals during the COVID-19 pandemic(17,27,29,30).

Findings from this study show that death anxiety is high among health professionals, specifically for doctors who treat COVID-19 in environments where protection equipment is not always available. Furthermore, the lack of professional and emotional preparation to face death rates in the magnitude experienced as a result of the pandemic has a negative impact in the mental wellbeing of physicians(18,31).

In our sample, the majority of care to patients with COVID-19 was provided by younger physicians, average age 47 with an average of 20 years of experience, who may lack the emotional maturity to deal with loss of life and threats to their own wellbeing. This is not surprising given directives from the Ministry of Health in most Latin American countries designed to minimize risk to older doctors who may have co-morbidity factors placing they at higher risk for death from a COVID-19 infection.

When we analyze the fear of death from the perspective of facing the physician’s own mortality, data from our sample suggest that doctors who did not treat patients with COVID-19 were more afraid of their own death than their counterparts who treated COVID-19 patients. While doctors who cared for patients with COVID-19 reported being afraid for the end of life for their patients, doctors who did not treat those patients were more afraid of their own mortality. These findings are similar to those in previous studies (31).

Sample respondents presented an increase in fear associated with death in all the evaluated subcategories showing that health professionals are vulnerable to different mental health problems (18,20,25,26). Our findings also indicate greater levels of fear of dying for others among doctors with more years of experience. Similar feelings were found among doctors who treated patients with COVID and in those who have been graduated for a longer time. It is hypothesized that this fear is the result of greater exposure to death either as a result of more years of practice or more intense exposure to death due to COVID-19.

Finding from this study show that death anxiety for oneself and for others is higher among doctors who treated patients with COVID 19 and in retired doctors. This finding suggests that medical personnel need to obtain emotional and spiritual support to carry out their duties during this time(20,27,30).

Results from this study indicate that despite the fact death is a natural and unavoidable medical professionals who experience the loss of a patient also experience feelings of impotence and guilt which as some researchers have suggested could trigger physical or psychological symptoms 26. Furthermore, these results seem to suggest that at this time and as a result ofSARS-CoV2, the ever changing morbidity and mortality rates related to the pandemic, and the saturation and overload of health systems fear and death anxiety are high among the studied population.

## Conclusion

Several researchers have suggested that medical personnel need to be prepared to address feelings resulting from the death of their patients and from the potential of their own demise (17,20,21,30). Health care facilities must do more to better prepare physicians at all levels to better address fear and anxiety about death during pandemics. Future research could focus on studying the impact of culture and race/ethnicity on the expression of fear and anxiety.

## Data Availability

Data available upon request.

https://drive.google.com/drive/folders/1gf1Aue9SIlL-DhpogWigVSof28zsR_I-?usp=sharing

## Notes

### Competing Interest Statement

The authors have declared no competing interest.

### Funding Statement

No external funding was received for this study.

### Author Declarations

Comite de Etica. Universidad Ricardo Palma.

### Summary of Updates

Author order.

